# Information and Communication Technology (ICT) in Medical Education: A Survey among Medical Students’ of Bangladesh

**DOI:** 10.1101/2021.04.30.21256392

**Authors:** Jannatul Ferdoush, Fatiha Tasmin Jeenia, IkramUllah Khan, Tasfia Momtaj Chowdhury, Zahangir Alam, Md. Masud Morshed, Abhijit Chowdhury, Kohinoor Parveen, Maliha Ata, Rajat Sanker Roy Biswas

## Abstract

**Background:** Bangladesh is declared as Digital Bangladesh however medical students are least acquainted with the necessary ICT knowledge in the context of medical education. We have conducted a survey to know about Digital Equipment Ownership, therefore, carried out a self-reported assessment of knowledge and Utilization of ICT in relation to educational and clinical development.

**Materials and Methods:** A cross-sectional, multi-centered questionnaire survey was conducted among the medical students during the period of October 2019.

**Result & Discussions:** In total 467 medical students responded to the questionnaire in which 92% of the students owned a Google android smart phone and accessed the internet on their devices. 69% student has (1-5) medical related apps in their device and drug formulary apps most commonly used 43%. 59% of students have gained their present computer knowledge by via self-learning. For study work with fellow student, 90% student preferred Facebook group, Whats app and Viber. While there was a range of variation in the usage of non-academic search engines 72% Google and no usages were found for more dedicated academic services such as PubMed and Medscape. According to 74% of students, inadequate integration of ICT knowledge into their syllabus culminated in insufficient future professional skills. All students were keen for a Basic ICT learning program at the beginning of their medical courses.

**Conclusion:** The study revealed that to prepare medical students for using ICT in their academic work adequately, introducing ICT training at the initial stage of the undergraduate program and designing the curriculum to develop a multiplicity of skills is essential in addition to an integrated teaching method.

## Introduction

Use of Information & Communication Technology (ICT) has gained prominence in education, notably for medical education within the developed world [1]. The web may be a nice resource of knowledge and medical students can’t be unbroken removed from it. The move to include IT into undergraduate medical curriculum was initiated over a decade ago, discouragingly very minute evolution had been created toward meeting this goal, despite the fact that the requirement for this has being a lot of compelling. Medical undergraduates use computers oft for easy tasks, which cannot contribute to the event of data acquisition abilities [2]. Limited knowledge of ICT in learning context didn’t incorporate ICT into their educational purpose. Computer-assisted learning for medical undergraduates in management of various diseases was reported. [3-5]. Current guidelines of International standards for basic medical education powerfully support these initiatives [6-8]. Studies have systematically shown that using computers for learning develops problem solving skills [9] of medical students and improve performance at Multiple choice questions (MCQ), Objective structures clinical examination (OSCE) and Written assessments [10-11].

Instruction and engagement of medical students towards educational activities profoundly influenced by educators advanced readiness. One of the largest challenges in education nowadays is that academics communicate a noncurrent language from the pre-digital age [12]. This becomes a retardant after they teach a generation that speaks a wholly new language. Because of progression of medical education and Health care services framework overall it is important to stay up with the latest about data that is going on the world. There is enormous information we get in a single tick. What is significant isn’t simply essential information on the best way to utilize ICT yet in addition computerized proficiency, which incorporates the capacity to adjust to new difficulties and conditions in a quickly changing advanced world. This comprises abilities for basic facts recovery, information handling and the capacity to exploit the decent variety of computerized media.

The advancement in smart phone and web access alongside the expanding utilization of social media and free online courses and learning materials are altering the manner in which undergraduate learn, convey and work together. Smart phone serve as a source of anywhere resources [13] and upgrade educational and clinical activities, with the possibility to improve patient care. Increasing level of smart phone ownership and various medical related apps removes barriers [14] of adopting new technologies with immediate access of health related information leads to improve decision-making, reduced medical related errors.

Medical professionals now can hold text books, drug formularies, and disease guidelines on their cell phone [15-16] which updates regularly. However the employment of the web or smart phones can not be accustomed draw general conclusions regarding student’s ICT competencies and readiness [17]. Studies have shown that students with the lack of necessary skills to use computer-based learning platforms efficiently fall behind. [18].

Medical students of Bangladesh have high interest for the utilization of ICT for their studies and should be included during the initial stage of medical curriculum. It’s unfortunate that courses been educated at medical colleges don’t seem to be better preparing medical students to use ICT for their studies. While designing a curriculum, it’s vital to understand the expectation of students in traditional classroom environment with regards to their capabilities and resources. Such Information thus helps develop fundamental teaching and learning activities that enhance interaction and encourage students.[19]

To know the ground of medical students concerning ICT, we have conducted a survey to get a self-reported assessment on the usage of equipment, familiarity with software system, and the use of ICT for Educational and Clinical development.

## Materials and Methods

A cross sectional, multi-centered questionnaire survey was conducted among the 4th and 5th year medical student of Chittagong Medical College, BGC Trust Medical College, Army Medical College Chittagong, Bangladesh, during the month of October 2019. The questionnaire was constructed to explore the ownership of digital equipment, a self-reported assessment of usage of digital equipment, familiarity with software, use of ICT for educational and clinical development and need of students that preferred a formal ICT training program. The questionnaire was adopted from previous study [20-22].The questionnaire was tested in a small group of student to judge their understanding of the meaning of the questions. Later questionnaire was provided to the entire student who attended the particular class of the studied medical college. The students were asked to complete and return the questionnaire to the department of Pharmacology of respective medical college.

## Result

A total of 467 medical students answered the questionnaire. The male to female split was 44% (n=186) and 56% (n=281) respectively.

Table I represents that 92% (429/467) medical students owned a Google android platform smart phone. Majority of the students 89% (415/467) accessed the internet with their smart phones.

**Table I:**
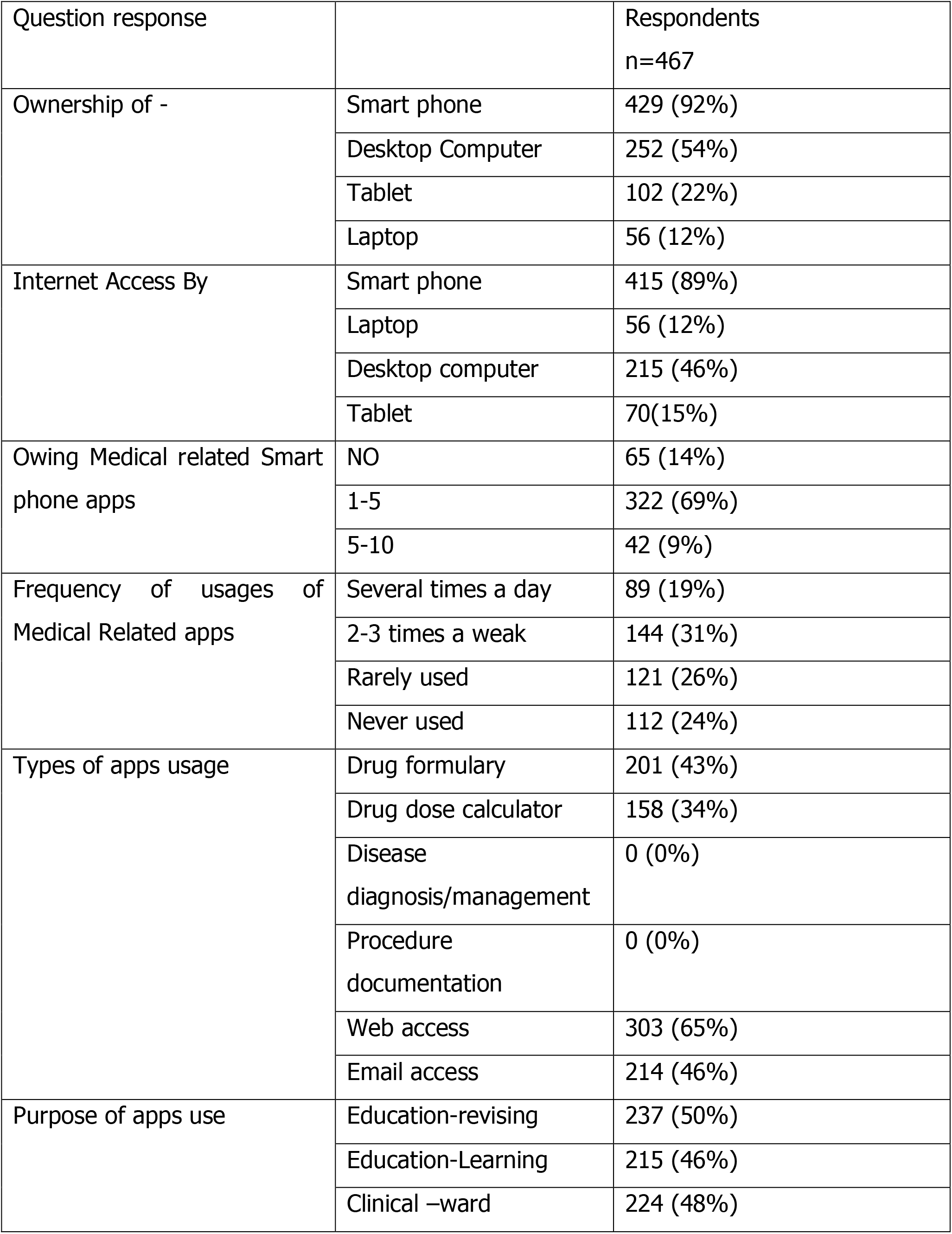
Medical student’s digital equipment ownership and usage.

69% (322/467) Students had at least 1-5 apps in their device, drug formulary apps most commonly used 43% (201/467), procedure and case documentation scored 0. Usages of apps for educational purposes were revision and learning equated to 50% (237/467) and 46% (215/467) respectively.

TABLE II represents that only 23% (107/467) students had gained their present computer knowledge through formal training programs and self-learning by 59% (276/467) students. 57% (266/467) and 21%(98/249) students mentioned that they know word processing and MS power point respectively while fewer students 7% (32/427) were familiar with spreadsheets (Table II).

**Table II:**
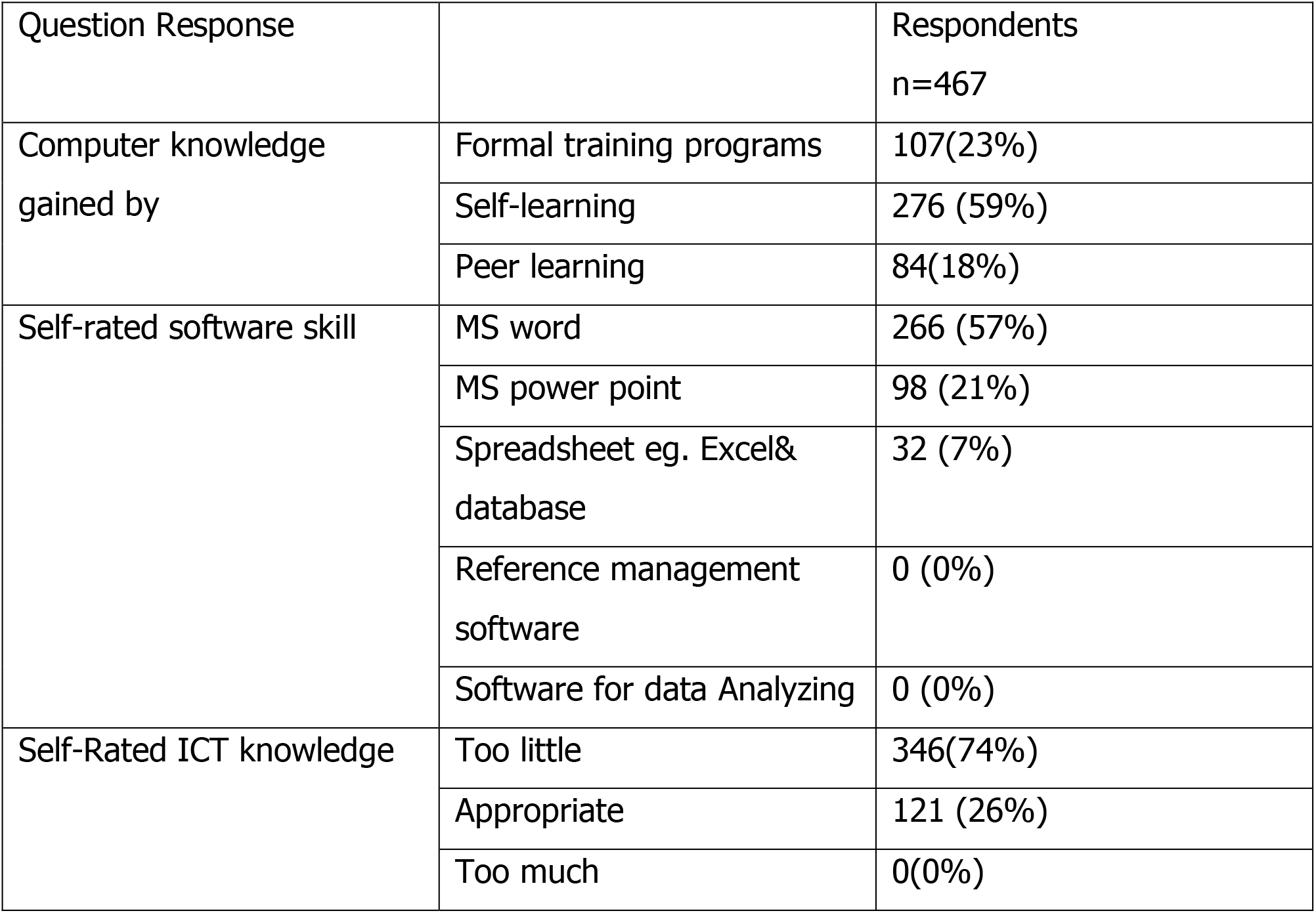
Medical Students Self assessed ICT knowledge and skill.

Table III represented that almost all students had their own e-mail address, of them only 55% (256) checked emails at least once a week. Students had a range of variation in the usage of non-academic search engines such as Google 72%(336/467)), narrow search engines such as Google Scholar 7% (33/467), no usages, howsoever was found for more dedicated academic services such as PubMed and Medscape. 53%(248/467) of the students used social networks such as Facebook in relation to their study whereas only 1% use professional networks and micro blogging such as LinkedIn and Twitter.

**Table III:**
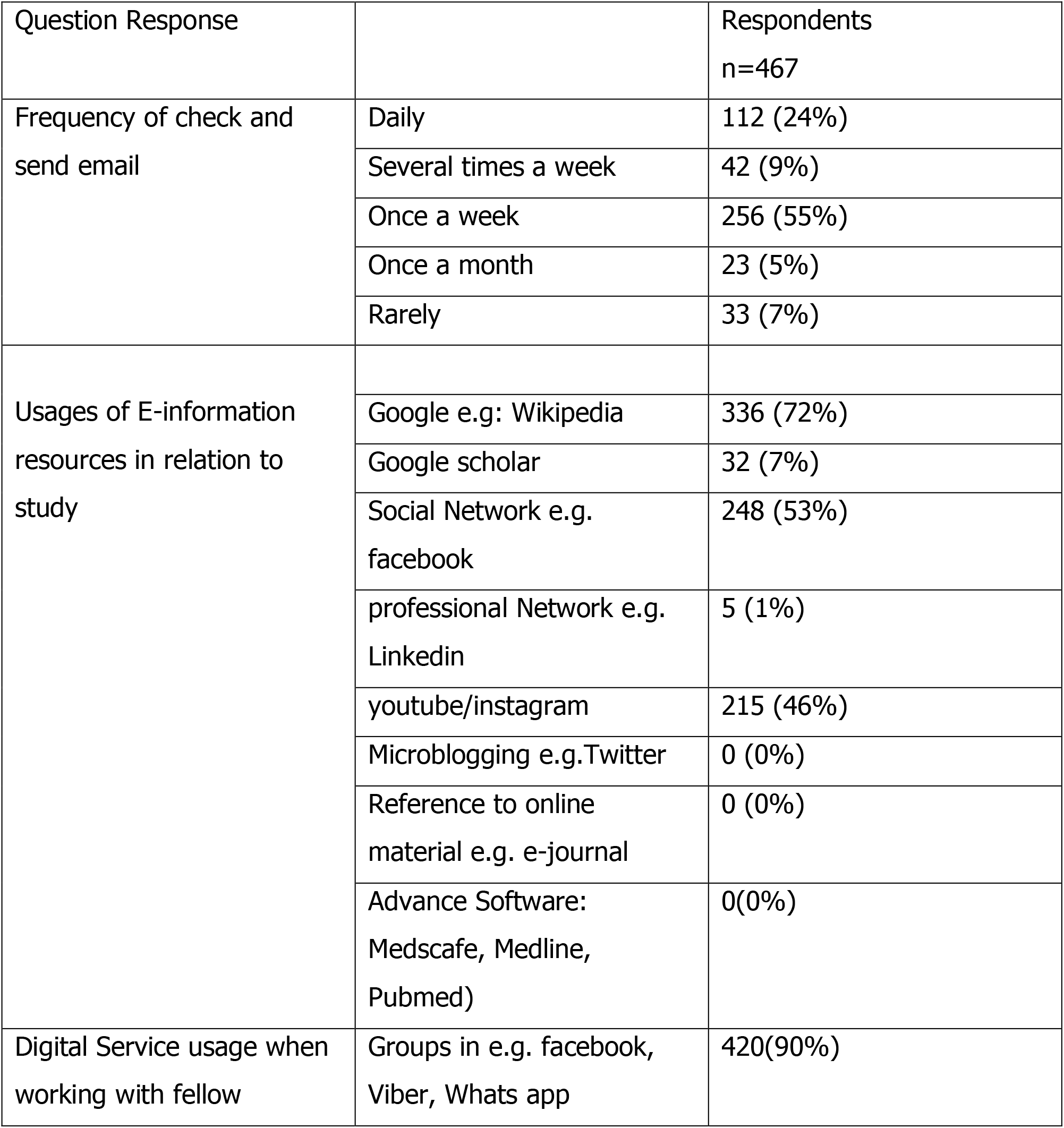
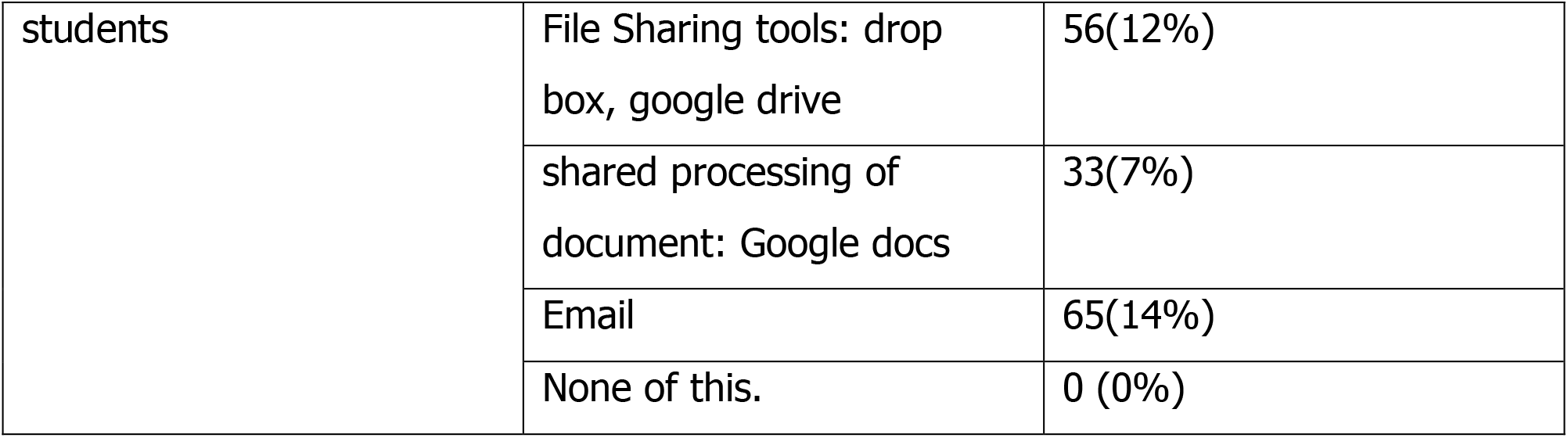
Usage of ICT in contest to medical education.

For study work with fellow students a combination of various digital platforms are used (table III), i.e. 90% (420/467) preferred group in Facebook, Viber, Whats app. Very few students 14%(65/467) preferred email as a way of communicating with fellow students. None were aware with reference management software and data analyzing software (Table III). Knowledge regarding ICT integrated to their educations is too little mentioned by 74% (346/467) undergraduates that were not satisfactory to provide them with the basic competency for their future profession.

**Table IV** represents that majority of the students (90%) were keen for a basic ICT learning program at the 1st year of their medical courses. 100% of undergraduates preferred to be trained on the searching and using medical resources on internet, Microsoft excel and statistical analysis software (Table IV).

**Table IV:** Needs of students that preferred a formal ICT program at faculty.

| Question Response |  | Respondents<br>n=467 |
| --- | --- | --- |
| Formal training on | Word processing | 154 (33%) |
|  | Power point presentation | 256 (55)% |
|  | Spreadsheet e.g MS excel | 467 (100%) |
|  | Statistical Analysis Software | 467(100%) |
|  | Searching and using medical resources on internet | 467 (100%) |
| Timing of training program | 1<sup>st</sup> year | 420 (90%) |
|  | 2<sup>nd</sup> year | 33 (7%) |
|  | 3<sup>rd</sup> year | 14(3%) |

## Discussion

To develop essential knowledge, skills, and ethics for effective and safe prescribing, ICT provides an expanse for medical students and health care professionals alike. Therefore, it’s vital to understand the patterns of usages of the digital equipment by students to develop acceptable learning materials and activities for delivery on digital platforms. This is a survey of the utilization of digital equipment and a self-reported assessment of ICT knowledge and skills of undergraduates’ medical students of Bangladesh. Of the medical students surveyed 92% (429/467) owned a Google android smart phone. Our results do collaborate with the findings of Payne FFB et al. study, where their majority of students 79.0% (248/467) owned smart phones and medical apps on their smart phone. [21]. Students largely owned desktop computers (54%), whereas only a minority (12%) of students had a laptop. The majority of the students (89%) report accessing the internet with their smart phones, 15% (70/467) with their tablets, and 46% (215/467) do so with computers. This finding indicates that those high levels of digital equipment usage by students will moderately be expected to use on-line material and participate in digital activities.

But once it involves the use of medical apps, 14%(65/467) of students haven’t any medical-related apps, only 69% (322/467) use 1-5 medical apps and only 9% (42/467) of the students use more than that. Only 19% (89/467) of them used medical apps several times a day. The types of apps use varied - drug formulary app (43%) drug dose calculator (34%), and web (65%) and email access (46%). Apps used for procedure/case documentation scored zero, possibly owing to the dearth of apps presently available in this area. Thus, the development of smart phone apps, in the context of Bangladesh, related to disease diagnosis/management, drug reference, and disease guidelines ought to be explored in future research. Desari k et al [23] reported 60% using medical apps for clinical activities and 47% for academic activities. In our study, 50% (237/467) students use medical apps for educational purposes-revision and learning, and 46% (215/467) using apps for clinical ward surroundings. According to another study conducted by Chowdhury NS et al, 35.7% of students use the internet for email & browsing; On one hand, 28.3% of students use the laptop solely for non-educational purposes like entertainment while on the other hand, 28% use the computer for educational purposes.[29]

Although our results showed that students had gained their present computer knowledge through formal training programs (23%), self-learning (59%) or by peer learning (18%), fundamental training of the students is also a significant predictor of computer literacy. Our finding is almost similar to the Ranasinghe et al study where 64.1% of students had formal training, 63.0% by self-learning, and 49.2% by peer learning. [22].

To obtain the benefit of a digitally supported educational environment students should be aware of the benefit of the software packages commonly used for educational activities. 57% (266/467) and 21% (98/467) of students mentioned that they know word processing and MS PowerPoint respectively whereas fewer 7% (32/467) were aware of spreadsheets. None of the students are aware of reference management software. Almost all of the students mentioned that they never used software for statistical analysis in reference to their studies. Similar findings from other resource-poor settings wherever the best levels of competency in areas were for Email, web, and File management. For alternative skills like data processing, most respondents mentioned low levels of a competency [24].

On the other hand, in developed countries nearly opposite findings are seen compared to ours. A study conducted among medical undergraduates in the United Kingdom revealed that they were aware of data processing (96%), spreadsheets (59%) and reference programs (23%). it’s shocking that digital natives don’t use tools like these. Therefore, they have got to think how institutional activities will be planned consistently to develop digital competences.

Knowledge regarding ICT integrated to their educations is too little to develop the required future professional competency mentioned by 74% (346/467) undergraduates in the present study. Similar findings in Woreta et al. stated that 44.0% of students rated themselves as less competent or a beginner of basic IT skill. [25] In Chowdhury NS et al Study, 61.8% of students thought they need average knowledge on computers, 8.1% of students mentioned they still don’t have the expertise to control computers, despite a number (11.2%) of them considering themselves to be skilled users.[26]. It is attention-grabbing that almost all of the students who participated during this survey also be thought-about to belong to the generation of digital natives.

It is necessary to students have the ability to obtain and use on-line information [20]. Nowadays, the information market is so congested that students need to know how to scrutinize authentic resources from this market. Almost all students owned e-mail addresses of them only 24% (112/467) checked emails daily. Students have multiple search choices once looking for information in regard to their study; a large proportion (72%) use non-academic wide search engines like Google, whereas a low proportion (7.0%) use narrow search engines like Google Scholar. Unlike the Vanozzi & Bridgestock study where 55-65% of European students use dedicated educational services like PubMed and Library search engines, none used such services in our study. Perhaps, It may be due to a lack of knowledge of how digital services can be used academically in contrast to personal use.[28].

In our study, 90% (420/467) of students preferred group in Facebook messenger, Viber for sharing information with fellows. Only a few students (14%) liked email as the way of communication as the delay in response makes it frustrating. Since our medical curriculum is yet to make any approaches in teaching and learning for the medical students, it has thus failed to accommodate students’ preferences in communication and ways of learning.

In the present study students opined that basic ICT learning program should be at the 1st year of their medical courses. The introduction of formal computer literacy courses to the first-year undergraduate medical program will produce a standard level of competency in order to use digital services effectively in their studies as well as meet their future professional needs.[27] Such skills include the ability to search a bibliographic database to obtain current practice guidelines, scientific information, participation in web conferences, and online teaching programs. However, such implementation would necessitate the overcoming of barriers like the finding of adequate time allocations from the busy undergraduate medical curriculum and also the lack of adequate resources at medical college for ICT training.

The typical learning environments in our medical college are mainly text-based, presumably mediated via a PowerPoint presentation. This is often an old-fashioned way of using technology and may be seen as a parallel to using a computer and word processor as a typewriter. However, the benefits of multimedia-based content delivery are seldom thought-about. And due to misalignment between student’s and teacher’s expectations of relevant teaching methods, the students are comparatively less familiar with dedicated educational tools and use social media less in regard to their studies compared to outside their studies.

It should be taken into account whether social media is perhaps relevant to be included within the educational activities. One example is the use of LinkedIn, which can be a platform for job opportunities and networking. Teachers should integrate ICT in teaching and learning activities such as using visual media, providing feedback, and creating environments that enable student-mediated approach to basic knowledge, materials, and the way to seek it out.

The present study has many limitations. We tend to fail to study the consequences of many alternative factors that would be related to ICT learning; whether it was absolutely an economic constraint or lack of interest that precluded digital equipment ownership and learning. Factors known throughout the current study can be used to improve ICT usage among medical undergraduates in Bangladesh and other developing countries with similar student populations and circumstances.

## Conclusion

The majority of the students claimed that the incorporation of ICT into their medical education is too inadequate to offer skills for their future profession. Although most students owned digital equipment, from Smart phones to laptops, had internet access and basic software knowledge, fewer familiarized with digital services concerning their academics. Hence, introducing ICT at the initial stage of the undergraduate program and redesigning the curriculum to encourage the usage of ICT in their medical studies is integral to the diversification of skills of students in addition to an integrated teaching method.

## Data Availability

It was a questionnaire survey among 4th and 5th year medical students of bangladesh.

